# Leveraging of SARS-CoV-2 PCR cycle thresholds values (Ct) to forecast COVID-19 trends

**DOI:** 10.1101/2021.07.17.21260679

**Authors:** Nicolas Yin, Simon Dellicour, Valery Daubie, Nicolas Franco, Magali Wautier, Christel Faes, Dieter Van Cauteren, Liv Nymark, Niel Hens, Marius Gilbert, Marie Hallin, Olivier Vandenberg

**Affiliations:** Department of Microbiology, Laboratoire Hospitalier Universtaire de Bruxelles – Universitair Laboratorium Brussel (LHUB-ULB), Université Libre de Bruxelles (ULB), Brussels, Belgium; Spatial Epidemiology Lab (SpELL), Université Libre de Bruxelles, Bruxelles, Belgium; Department of Microbiology, Immunology and Transplantation, Division of Clinical and Epidemiological Virology, Rega Institute, KU Leuven, Leuven, Belgium; Namur Centre for Complex Systems (naXys) & Department of Mathematics, University of Namur, Namur, Belgium; Interuniversity Institute for Biostatistics and statistical Bioinformatics (I-BioStat), Data Science Institute, Hasselt University (UHasselt), Hasselt, Belgium; Scientific Directorate of Epidemiology and public health, Sciensano, Brussels, Belgium; Norwegian Institute of Public Health, Division of Infection Control and Environmental Health, Oslo, Norway; Department of Health Management and Health Economics, University of Oslo, Oslo, Norway; Centre for Health Economic Research and Modelling Infectious Diseases, Vaccine and Infectious Disease Institute, University of Antwerp, Antwerp, Belgium; Centre for Environmental Health and Occupational Health, School of Public Health, Université Libre de Bruxelles (ULB), Brussels, Belgium; Clinical Research and Innovation Unit, Laboratoire Hospitalier Universitaire de Bruxelles - Universitair Laboratorium Brussel (LHUB-ULB), Université Libre de Bruxelles (ULB), Brussels, Belgium; Division of Infection and Immunity, Faculty of Medical Sciences, University College London, London, United Kingdom

**Keywords:** COVID-19, SARS-CoV-2, forecast, epidemic trend, Ct values

## Abstract

**Introduction:** We assessed the usefulness of SARS-CoV-2 RT-PCR cycle thresholds (Ct) values trends produced by the LHUB-ULB (a consolidated microbiology laboratory located in Brussels, Belgium) for monitoring the epidemic’s dynamics at local and national levels and for improving forecasting models.

**Methods:** SARS-CoV-2 RT-PCR Ct values produced from April 1, 2020, to May 15, 2021, were compared with national COVID-19 confirmed cases notifications according to their geographical and time distribution. These Ct values were evaluated against both a phase diagram predicting the number of COVID-19 patients requiring intensive care and an age-structured model estimating COVID-19 prevalence in Belgium.

**Results:** Over 155,811 RT-PCR performed, 12,799 were positive and 7,910 Ct values were available for analysis. The 14-day median Ct values were negatively correlated with the 14-day mean daily positive tests with a lag of 17 days. In addition, the 14-day mean daily positive tests in LHUB-ULB were strongly correlated with the 14-day mean confirmed cases in the Brussels-Capital and in Belgium with coinciding start, peak and end of the different waves of the epidemic. Ct values decreased concurrently with the forecasted phase-shifts of the diagram. Similarly, the evolution of 14-day median Ct values was negatively correlated with daily estimated prevalence for all age-classes.

**Conclusion:** We provide preliminary evidence that trends of Ct values can help to both follow and predict the epidemic’s trajectory at local and national levels, underlining that consolidated microbiology laboratories can act as epidemic sensors as they gather data that are representative of the geographical area they serve.

## Introduction

The coronavirus disease 2019 (COVID-19) pandemic dramatically highlighted the central position of diagnostic testing, not only for the clinical management of infected individuals but also for surveillance purposes ^1^. The use of clinical microbiology laboratories (CMLs) data to survey the presence of specific microorganisms in a given population represents one of the most established public health surveillance tools of infectious diseases. In a previous study, we proved that influenza trends in Belgium may be estimated using laboratory data provided by a CML serving the wider Brussels-Capital Region area ^2^. Since the start of the COVID-19 pandemic, several authors have demonstrated that CMLs could represent the first step toward a global set of sensor networks for infectious diseases surveillance, where each one of the CMLs can be seen as a real-time sensor in its area within an interconnected, complex network ^1,3,4^. In this perspective, CMLs have become a cornerstone in the fight against SARS-CoV-2 infections due to their ability to process large amounts of samples in large geographic areas while using highly specialized diagnostic tests ^1,5^.

By reporting to Sciensano, the Belgian national public health research institute, the number of new positives among the tests conducted each day, CMLs share the data needed to estimate the effective reproduction number (R_t_) ^6,7^. However, the data represent the growth rate of positive tests and not the incidence of infection, which requires adjustments to account for changes in testing capacity, delay between infection and test report date, and conversion from prevalence to incidence. We previously showed that SARS-CoV-2 RT-PCR cycle threshold (Ct) values are different between populations, with lower Ct values – thus higher viral loads – for outpatients, likely to be recently infected and higher Ct values for inpatients ^8^. In a recent article, Hay *et al*. used the SARS-CoV-2 RT-PCR Ct values in a model to forecast epidemic’s trajectory ^9^. At the time of writing, RT-PCR assays are not standardised and the Ct values obtained using various PCR methods on various instruments in various laboratories using various sampling methods cannot be easily aggregated by surveillance systems. Sciensano recently encouraged laboratories to report their results using a semi-quantitative approach where a viral load below 10^3^ RNA copies/mL is considered as “weak positive” ^10^. Sciensano’s primary goal was to approach the actual infectiousness of patients with persistent positive RT-PCR. Therefore, the semi-quantitative dimension of positive test results is not used by surveillance systems yet.

Besides the difficulty of making use of all the data provided by CMLs in real time, public health authorities also face the challenge of making decisions, as the constantly evolving situation requires permanent adaptation ^11^. In this perspective, various predictive models have been developed to support policy makers ^12–15^. To improve and facilitate the decision-making process, Hens *et al*. developed a phase portrait to monitor the epidemic allowing a real-time assessment of whether intervention measures are needed to keep hospital capacity under control ^16^. Nevertheless, such supportive decision tools are often designed at the national level instead of the hospital level where, during the pandemic, hospital managers needed support to forecast the cancellation and reintroduction of a series of medical activities, such as the surgical care program, or the number of COVID-related ICU beds ^17^. Thanks to the huge amount of data they collect on a daily basis, CMLs could also help the hospital structures they serve to anticipate the evolution of the epidemic and forecast their hospitalisation and medical activities.

The objectives of this study were: (1) to verify the accuracy of using of SARS-CoV-2 PCR Ct values trends in a single CML to monitor the dynamics of the epidemic; (2) to determine the added-value of using these data as an additional advanced information for scenario analysis, in relation to a phase diagram and an age-structured compartmental model, both developed to follow the path of the Belgian COVID-19 epidemic ^14,15^.

## Methods

The “Laboratoire Hospitalier Universitaire de Bruxelles - Universitair Laboratorium Brussel” (LHUB-ULB) is a merged clinical laboratory serving five university hospitals located in the Brussels-Capital region in Belgium ^8^. All the SARS-CoV-2 PCR results produced between April 1, 2020, and May 15, 2021, by the LHUB-ULB were extracted anonymously from its laboratory information system. The data collected were patients’ postal code, age, qualitative PCR results, Ct values, instruments on which PCR were performed, and sampling dates. National Belgian data were extracted from the “total number of tests by date” and the “confirmed cases by date, province, age and sex” public dataset available on the Sciensano website on May 27, 2021. These datasets contain the total number of tests, the number of positive tests per day, and the confirmed number of cases per day and province. To analyse trends and minimize day-to-day and holiday-related fluctuations, we computed mean daily positive tests and cases, and median and mean Ct values from May 1, 2020 to May 15, 2021, using a backward sliding window of 14 days (hereafter referred as “14-day mean positive tests/cases” and “14-day median/mean Ct values”).

To follow the trends of Ct values variation during the study period, only the SARS-CoV-2 PCR results on nasopharyngeal swabs (NPS) obtained using the *m*2000 RealTi*m*e SARS-CoV-2 assay (Abbott Molecular, USA) were considered, this assay being the only one used by our laboratory during the entire period of interest. As detection of both targeted genes (RdRp and N) was performed using the same fluorophore, the Ct values of this assay were observed up to 32 cycles and were not comparable with Ct values of other RT-PCR assays. Ct values were plotted against a standard calibration curve provided by the Belgian NRC to obtain the semi-quantitative results recommended by Sciensano ^10^. Accordingly, results with a Ct > 22.3 were considered as “weak positive” (viral load < 10^3^ RNA copies/mL). Correlations between 14-day median/mean Ct values and daily mean positive tests were calculated using Spearman’s *r*_S_ rank correlation coefficient. This correlation was performed with shifts of 0 to 30 days in the median and mean Ct values, to determine the shift with the highest *r*_*S*_ between the daily mean number of positive tests and Ct values. To test their validity as a source for COVID-19 surveillance, LHUB-UB’s data were also compared with all COVID-19 confirmed case notifications according to geographical coverage and time distribution.

We used phase diagrams depicting the evolution of COVID-19 hospitalisations in Belgium to compare these trends with the evolution of Ct values measures through time ^16^. These diagrams were developed to predict the number of COVID-19 patients requiring intensive care by considering the 7-day mean new hospitalisations and the daily ratio of the past 14-day new hospitalisations. For each combination, the total number of hospitalisations is projected for a horizon of 14 days, from which the number of patients requiring intensive care is predicted based on the distribution of the time spent in an intensive care unit (ICU). The hospital contingency plan in Belgium consists of five different phases (phases 0, 1A and 1B, 2A and 2B), incrementing COVID-19 related ICU beds capacities. Within this scheme, the total number of patients in ICU moves from 2001 to 2821, consequently yielding a gradual decrease in non-COVID-19 ICU capacity ^16^. The hospital and future COVID-related ICU load is thus depicted from green to red: the green region can be considered a “safe zone” in which the number of new hospitalisations is limited with a decrease (growth < 1) or a limited increase (growth > 1) and associated with a limited number of COVID-19 patients at ICU (first part of Phase 0); the yellow region, a region of increased vigilance (second part of Phase 0). The orange (Phases 1A & 1B) and red (Phases 2A & 2B) regions are “high impact” and “no-go” zones, in which non-COVID-19 care decreases substantially and additional capacity for COVID-19 needs to be provided for.

A comparison between the evolution of 14-day median Ct values by age classes and the daily estimated Belgian COVID-19 prevalence for theses age classes has been performed using a model of deterministic continuous age-structured compartmental model (extended SEIR-type) integrating social contact data and calibrated on hospitalisations and deaths incidence data as well as serological studies ^15^. The prevalence was estimated for the following age classes in years: 0-24, 25-44, 45-64, 65-74 and 75+ as the proportion of the sum of the infected compartments (exposed, asymptomatic, presymptomatic, symptomatic and hospitalised individuals) compared to the total size of the age class, with a 90% confidence interval estimated by Bayesian analysis. This method aims to provide a reliable comparison with the spreading of COVID-19 in Belgium among age classes since the number of RT-PCR positive tests are known to be biased over time due to testing policy changes, especially regarding the youngest and oldest classes.

Data from all sources were collected retrospectively and anonymously before analysis from a routine surveillance perspective. Ethics approval was granted by the Ethics Committee of the Saint-Pierre University Hospital. No written informed consent was collected.

## Results

From April 1, 2020, to May 15, 2021, a total of 155,049 SARS-CoV-2 RT-PCR were performed in the LHUB-ULB and resulted in 12,771 positive results of which 7,906 Cts were analysed. A peak of LHUB-ULB 14-day mean daily positive tests was reached during the Belgian second wave on October 28, 2020 (n = 153.6, Fig. 1). Beforehand, a lower peak was reached during the summer on August 22, 2020 (n = 24.4). In both cases, these peaks were preceded by a drastic decrease in the 14-day median Ct values reaching local minima respectively 16 days before (13.12 on October 12, 2020) and 12 days before (12.76 on August 10, 2020). Ct values were negatively correlated with the number of LHUB-ULB positive tests, with a maximum reached for the correlation between the 14-day median Ct values with a lag of 17 days and the 14-day mean positive tests (*r*_*S*_ = - 0.836), as well as between the 14-day mean Ct values with a lag of 19 days and the 14-day mean positive tests (*r*_*S*_ = −0.834).

**Figure 1.**
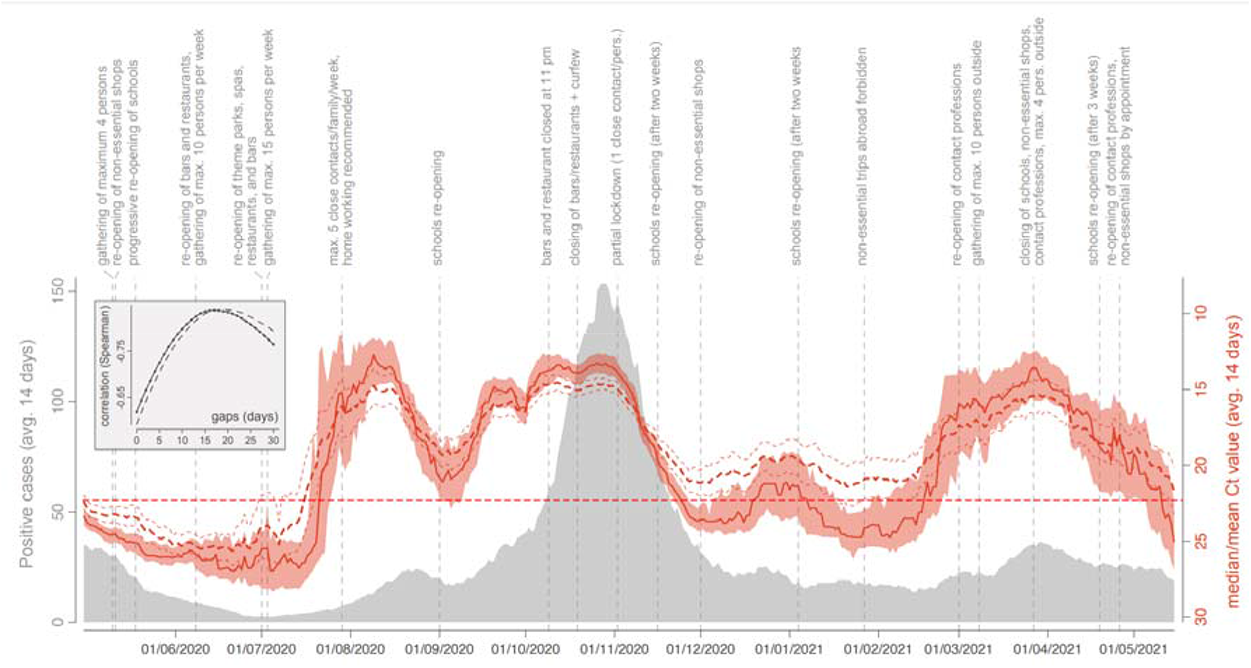
COVID-19 trends in a laboratory perspective: evolution of the 14-day median (solid red curve) and mean (dashed red curve) Ct values obtained from April 1, 2020, to May 15, 2021 (LHUB-ULB, Brussels Region, Belgium). Reddish areas surrounding the solid red curve indicate the 95% confidence interval (CI) associated with median Ct values, and the thinner red dashed lines indicate the 95% CI associated with mean Ct values. The horizontal dashed line refers to the threshold value of 22.3. Those curves are superimposed on the evolution of the number of COVID-19 positive cases identified at the LHUB-ULB. As detailed in the text, median and mean Ct values, as well as COVID-19 positive cases, were all computed using a backward 14-day sliding window. In the embedded box, we report the relationship between (i) the Spearman correlation between daily estimates of median (solid curve) and mean (dashed curve) Ct values and the number of COVID-19 positive cases (LHUB-ULB) and (ii) the time gap considered between the Ct measures and the number of positive cases. On top of the graph, we also report key dates of the Belgian epidemic.

During the same period, a total of 1,381,393 tests were performed across the Brussels Region and 13,219,135 tests across the whole of Belgian territory, of which respectively 142,562 and 1,131,719 tests were positive. Overall, LHUB-ULB performed respectively 8.96% (12,771/142,562) and 1.13% (12,771/1,131,719) of all positive tests reported in the Brussels Region and at the national level. Figure 2 shows the geographical distribution by postal code of the confirmed COVID-19 cases notified by the LHUB-ULB to Sciensano and the LHUB-ULB’s representativeness in the COVID-19 notification. Beside the Brussels-Capital Region, which concentrated most of the tests produced by the LHUB-ULB, its service area extended to several municipalities in Walloon and Flemish Region with, for some of them, about 5% of all notifications. Overall, the number of positive tests produced by the LHUB-ULB showed a high correlation with the regional and national trends of the incidence of COVID-19 notifications with coinciding start, peak and end of the different waves of the epidemic. The 14-day average number of positive tests in LHUB-ULB were strongly correlated with the 14-day average number of positive tests in the Brussels-Capital Region (*r*_S_ = 0.843) and in Belgium (*r*_S_ = 0.810) but also with the 14-day average confirmed cases in the Brussels-Capital Region (*r*_S_ = 0.832) and in the whole country (*r*_S_ = 0.804) (figure 3).

**Figure 2.**
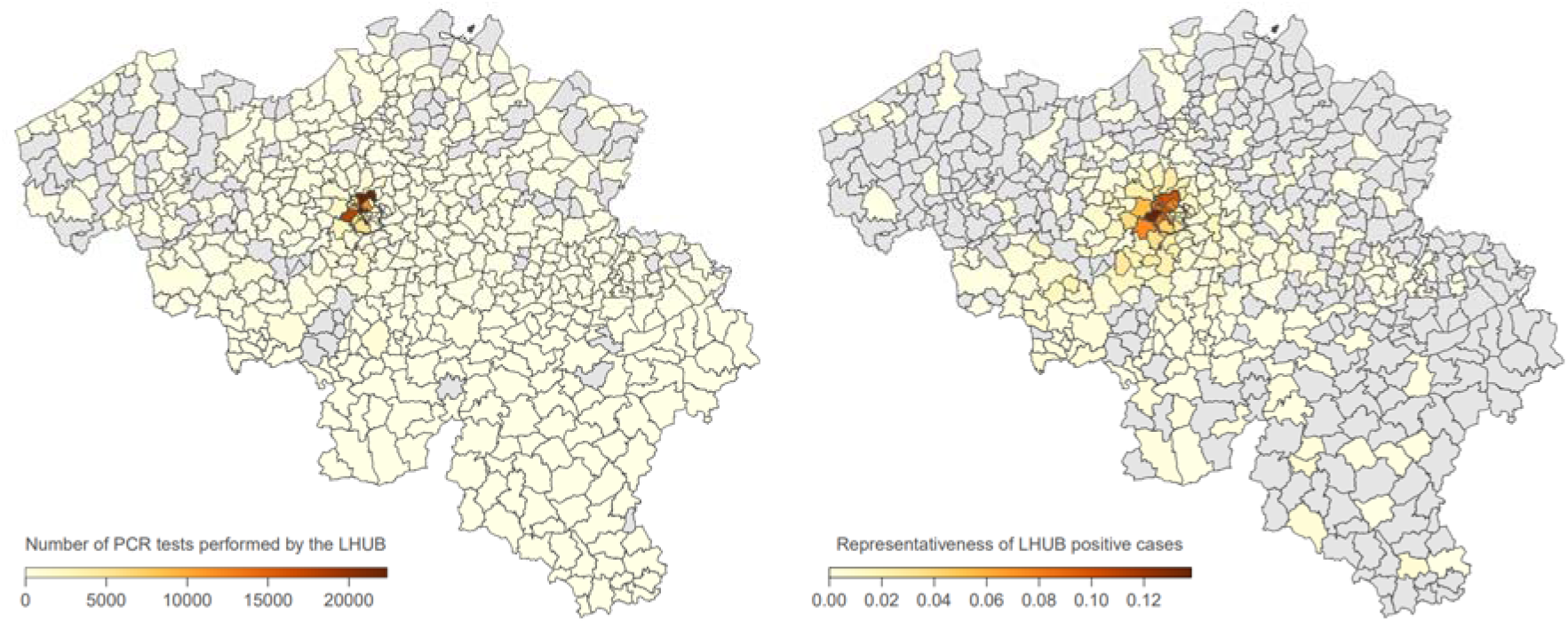
Mapping by municipality of SARS-CoV-2 RT-PCR tests performed by the LHUB-ULB since March 8, 2020, and of the proportion of positive cases detected by the LHUB-ULB compared to the overall number of positive cases detected in those areas. The Brussels Region is highlighted by a dark grey contour.

**Figure 3.**
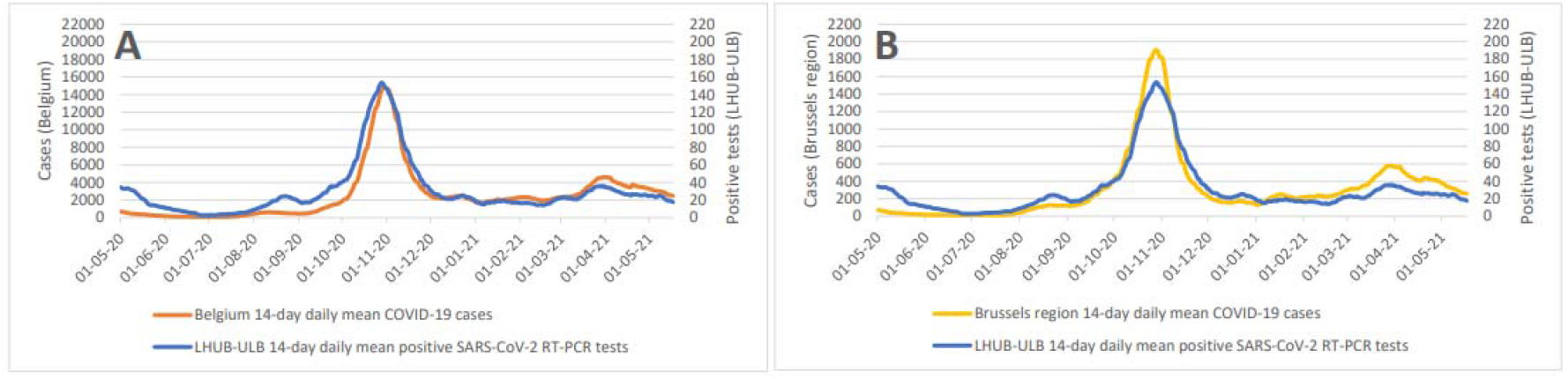
Compared evolution of 14-day mean number of positive SARS-CoV-2 RT-PCR tests (LHUB-ULB) with Belgium (A) and Brussels (B) 14-day mean COVID-19 cases.

In Figure 4, the 14-days median estimates of daily Ct values are plotted in a white to blue colour scale on the phase diagram introduced above, showing how Ct values decrease when the situation worsens (and vice versa) in trends making clockwise movements. Figure 4A shows the downward trend of the end of the first epidemic wave during which the growth in new hospitalisations progressively decreased to reach below 0%, a moment at which the number of new hospitalisations started to decline: The Ct values, low at the peak, increase when the number of new hospitalisations starts to decline. In Figure 4B, an upward trend was observed, leading to a small summer wave. As soon as both growth and hospitalisations passed from the green “safe zone” to the yellow region of “increased vigilance”, the Ct values started to decrease, concurrently crossing the threshold value of 22.3. The opposite effect was observed when the points fell in the green region. The second wave is visualised in Figure 4C, with a clear decrease and increase of the Ct values. Finally, Figure 4D corresponds to the third wave, with again the same pattern observed in the evolution of Ct values.

**Figure 4:**
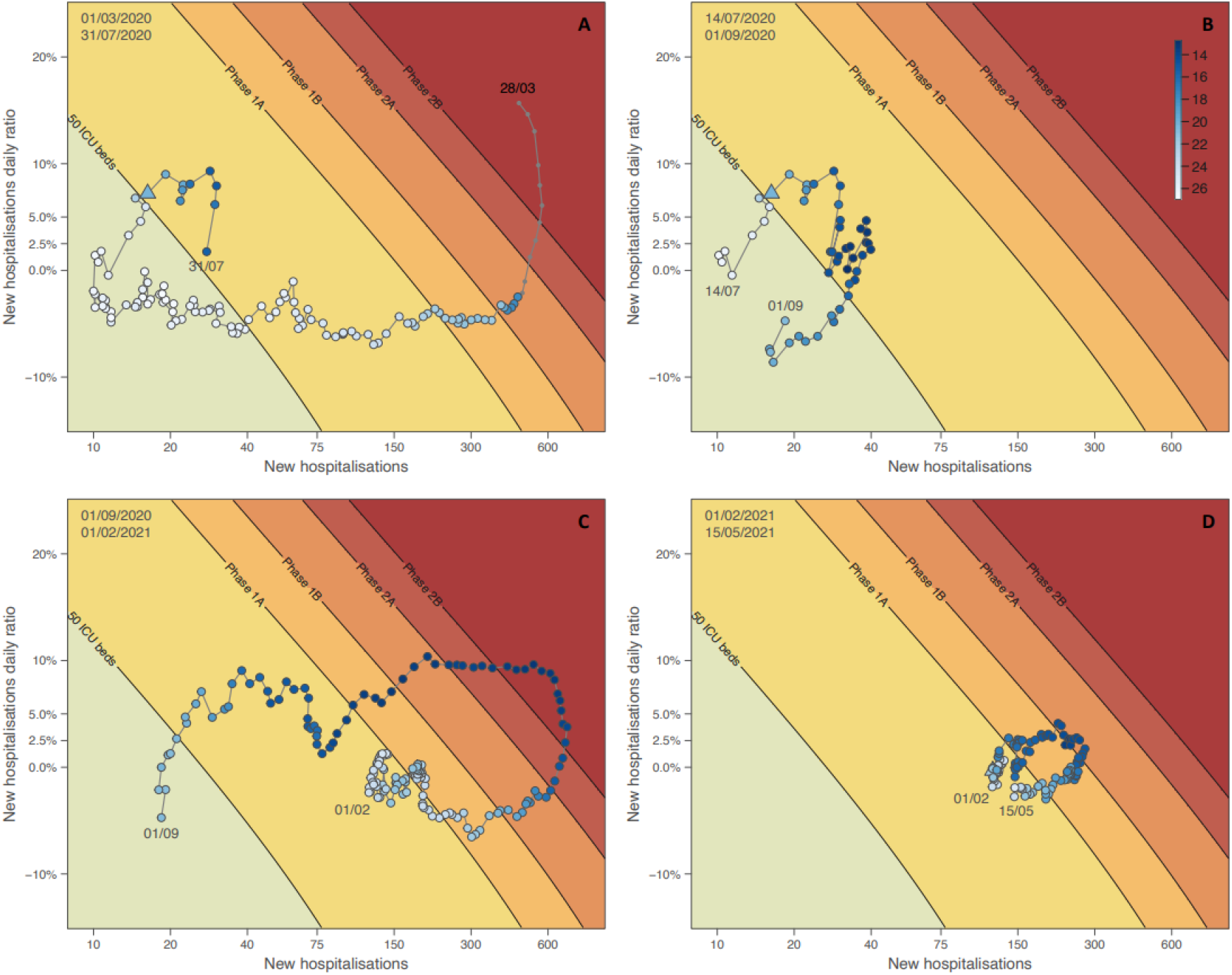
Phase diagram generated for different time periods: situation from March 1 to July 14, 2020 (A), situation from July 14 to September 1, 2020 (B), situation from September 1, 2020, to February 1, 2021 (C), situation from February 1 until May 15, 2021 (D). See the text for further detail on the principle of phase diagrams. Dots of the phase diagram are coloured according to 14-day median Ct values (thus computed using a backward sliding window of 14 days). Triangle symbols indicate the dates when Ct values crossed down the threshold value of 22.3.

In Figure 5, the median of daily Ct values for different age groups are compared to the daily estimated prevalence of those age groups. The overall behaviour of Ct values was almost similar for all age classes and was negatively correlated to the estimated prevalence.

**Figure 5:**
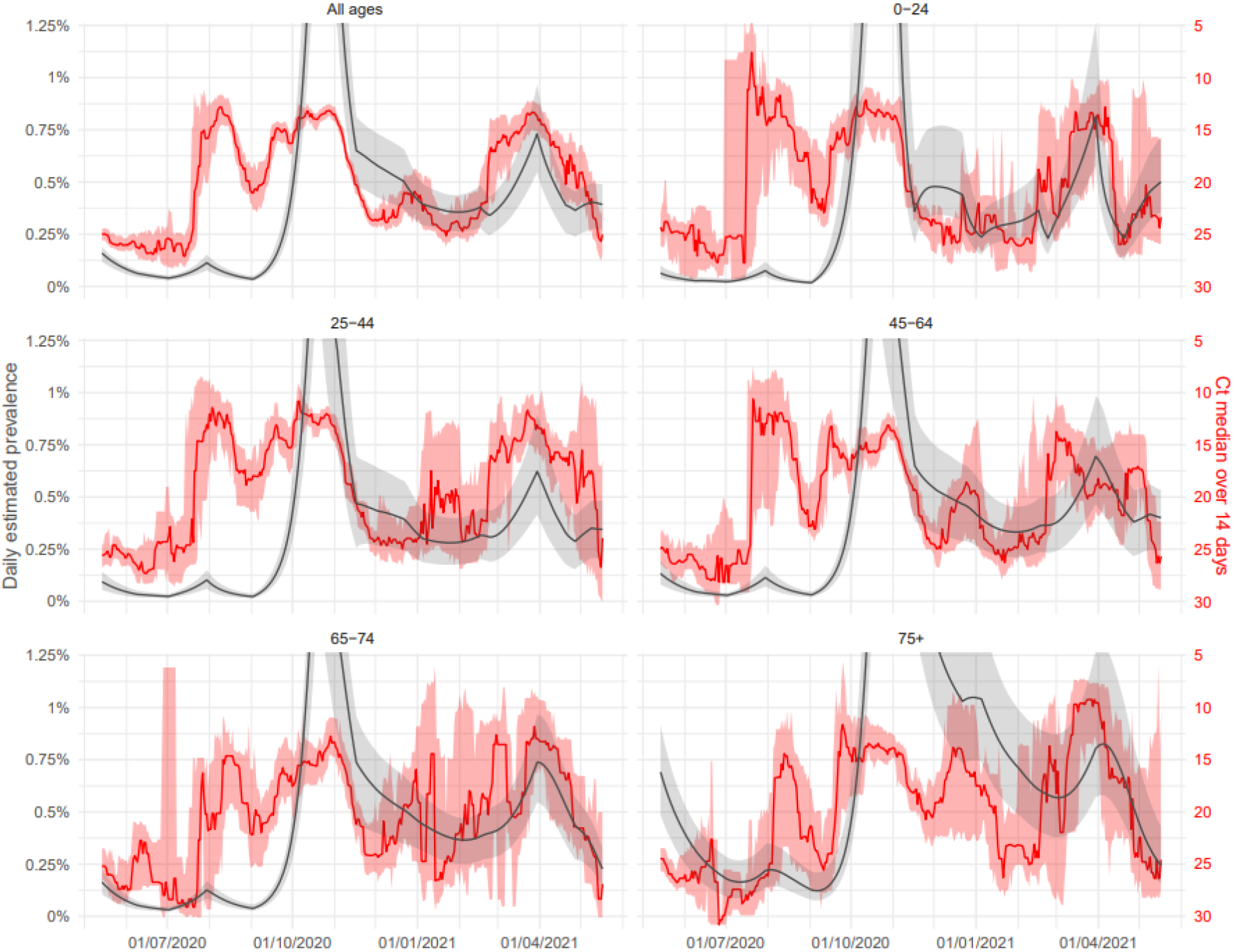
Ct evolution by age classes with comparison to the estimated prevalence: evolution of the daily 14-day median Ct values obtained using the Abbott RealTi*m*e SARS-CoV-2 assay from April 1, 2020, to May 15, 2021 (LHUB-ULB, Brussels region, Belgium). Reddish areas surrounding solid red curves indicate the 95% confidence interval (CI) obtained by bootstrapping method associated with daily estimates of median Ct values. Those curves are superimposed on the daily prevalence of COVID-19 infected people (proportion of infected versus Belgian demographic data) as estimated by the age-structured extended SEIR-type mathematical model. Solid grey curves indicate the 95% CI obtained by Bayesian analysis.

## Discussion

Estimating the likely number of infected patients during epidemics but also the dynamics of spreading in the population is crucial to carry out adequate testing and infection control measures. As large and accurate data providers, CMLs can adequately support hospital capacity planning by providing valuable real-time information about the incidence trends of the pandemic. This was already established by a previous study on influenza ^2^, but seems to be even more relevant in the context of a more severe disease like COVID-19, where hospital capacities are crucially challenged. Indeed, LHUB-ULB processed on its own 8.95% of the SARS-CoV-2 testing in the Brussels-Capital Region and was proved here to be representative not only of the region but to a certain extent, the whole country due its central geographic position in Belgium. A step further would be to capitalise on the ability of these CMLs to rapidly detect and communicate abnormal events such as sudden increase or emergence of variants of concern without the delay resulting from sending samples to central sequencing platforms. Thanks to the expertise gained in such data integration, UK scientists were able to rapidly share an early assessment of the variant Alpha’s (lineage B.1.1.7) genomic characteristics and associated clinical outcomes ^18^.

Complementarily, and providing an adequate standardisation under appropriate management and regulatory structures, “virtual” CMLs consolidation can also adequately support ongoing COVID-19 surveillance by connecting some or all the produced data to national public health surveillance systems. In the frame of the COVID-19 pandemic, Sciensano started to monitor on a daily basis the epidemiological situation of SARS-CoV-2 in the country through multiple surveillance systems including the “healthdata.be” platform aggregating all information from all CMLs located in Belgium ^19–21^. The added value of such a combined structure was already demonstrated for monitoring viral infections by the Infection Response Through Virus Genomics–ICONIC consortium in London ^22^.

Beyond the variation of the infectiousness over time, our results suggest that following the trend of SARS-CoV-2 RT-PCR Ct values could predict the epidemic trends. Recently infected patients are known to have higher viral load, thus higher infectiousness ^23^. A decrease in Ct values, linked to an increase of recently infected people is likely to favour spreading, and goes hand in hand with an increase in the total number of cases. By gathering enough comparable data using semi-quantitative results, our Ct values based surveillance systems could approach in real time the average level of viral load in the population, hence approach the current spreading of the virus before the increase of cases becomes apparent, while avoiding the recurrent problem of normalization. Predicting the shape and the size of the epidemic curve is not straightforward; and many parameters may influence it such as seasonality, infection control measures and population immunity level, to cite a few. The evolution of 14-day median Ct values was also tested against the daily estimated prevalence by age classes, and Ct values were similarly negatively correlated for all age classes, even we observed a shift by approximatively half a month for the 75+ which might be due to intergenerational transmission. However, a starting divergence was observed in May 2021, with Ct values increasing for the oldest classes while remaining low for the youngest one. This was related to a period of resumption of activities in Belgium such as reopening of schools, while older age-classes were progressively becoming protected through the vaccination campaign. The prevalence projections from the compartmental model followed the same trend. Hence, Ct values divergence by age classes could be a good indicator of a divergence in transmission in these age classes.

Following the trend of the Ct values might have helped the decision makers as demonstrated with the integration of the Ct values in a phase diagram predicting the number of COVID-19 patients requiring intensive care at a national level. For instance, in March 2021, after a long period of stagnation in the epidemic, the Belgian government decided to reopen close-contact professions and increased the number of people authorized to gather outside, at a time when Ct values were decreasing. This reopening was reversed a few weeks later due to the increase of cases underlining the untimely decision. During the summer 2020, the evolution of Ct values accurately followed the dynamic of the epidemic with an increase accompanying each decrease of the pressure on hospitals. But the shift between the initial diagnosis, the admission, and the length of stay for COVID-19 inpatients makes it harder to anticipate the trends in hospitalisation between October 2020 and March 2021 when the epidemic had no real break between peaks and the tension in hospital beds remained stable. Only future evolution after a real epidemic reflux could confirm the added value of following the Ct values to anticipate the phase shifts.

At the hospital level, being able to foresee epidemic dynamics could allow a greater ability to anticipate measures such as pre-admission screenings, isolation, postponement of non-urgent interventions, triage, and upscaling of human resources. In our study, each epidemic wave was preceded by a drastic decrease of Ct values, the median crossing back the Ct = 22.3 value threshold (i.e. the proportion of “weak positive” tests went below 50%), setting here an eventual easy-to-evaluate parameter at the local level. This threshold value of 22.3 was clearly crossed back concurrently with the passage of the number of new hospitalisations versus the new hospitalisations’ daily ratio from the green “safe zone” to the yellow region of “increased vigilance” in the phase diagram. Even if the setting described here should likely be adjusted before being transposed to other laboratories to take account of the specificity of their own patients (ratios inpatients/outpatients and symptomatic/asymptomatic), repeating this exercise with their own data could allow them to set up their own alarm threshold. Likewise, local and national surveillance systems should track the difference in the proportion of strong versus weak positive results to model the dynamics of the epidemic and thus to provide guidance for prevention measures as suggested by Hay *et al* ^9^.

A potential weakness of our data is that a limited part of the LHUB-ULB activity relies on ambulatory patients at the general practitioner level. Being able to reach this “non-hospitalised” population would likely increase the sensitivity of a surveillance system to weak signals when the epidemic begins in the community before affecting hospitals. However, the fact that overall behaviour of Ct values was almost similar for all age classes and was negatively correlated to the estimated prevalence in the compartment model indicates that our data capture the whole Belgian population to a sufficient extend. One could also argue that correlation between Ct value and actual viral load depends on many factors, such as sampling method, targeted genes, primers and probes, and possible mutations in targeted genes ^24^. Due to the absence of standardisation between SARS-CoV-2 RT-PCR assays, we only analysed Ct values obtained using one RT-PCR assay performed on nasopharyngeal swabs all along the studied period. We do believe that their number is sufficient to neutralize the effect of measurement bias. Furthermore, it has been discussed that some variants could exhibit an average higher viral load ^18,25,26^ which could directly impact observed trends in the overall evolution of Ct values. Nevertheless, this potentially higher viral load is likely to favour infectiousness and should not introduce a bias regarding epidemic surveillance.

In conclusion, this study established a correlation between the trends in the SARS-CoV-2 RT-PCR Ct values and the trends of the COVID-19 incidence a few days later. Following the dynamics of the average viral load could add a dimension in the surveillance of respiratory infectious diseases. Moreover, it underlines that the considerable amount of data daily collected by CMLs can play a key role at both local level and beyond, depending on the geographical area they serve. By gathering comparable laboratory data approaching the average viral load of respiratory viruses in the population, surveillance systems might be able to better follow epidemic dynamics, establish forecast models, capture weak signals, and thus anticipate uncontrolled spreading.

## Data Availability

The complete de-identified data set will be available upon request to for researchers whose proposed use of the data has been approved, for any purpose. If needed, requests will require the ethics committee approval of the Saint-Pierre University Hospital (Brussels, Belgium). Anonymized data are fully available on reasonable request from the corresponding author after approval by the hospital ethics committee. Data Availability Statement. R script and related files needed to run the analyses and generate Figures 1, 3, 4 and 5 presented in our study are all available at https://github.com/sdellicour/Ct_measures_LHUB.

## Contributors

NY, SD, VD, LN, MH, and OVDB designed the study. NY and SD managed the database. NY, SD, DVC, VD and NF did the statistical analyses. NY, VD, MW and MH validated the laboratory analyses on clinical samples. NY, SD, VD, NF, CF, MH, OVDB accessed and verified the data. NY, SD, NF, MH and OVDB wrote the paper. All authors had the opportunity to discuss the results and comment on the manuscript. All authors had full access to all the data in the study and had final responsibility for the decision to submit for publication.

## Declaration of interests

All authors declare no competing interests.

## Acknowledgments

We wish to thank the personnel of the LHUB-ULB for its daily technical assistance. The authors would like to thank Dirk Thielens for his appreciable assistance. This work is dedicated to the healthcare workers, the patients, and families affected by SARS-CoV-2. SD is supported by the Fonds National de la Recherche Scientifique (FNRS, Belgium). NF, CF and NH acknowledge support from the European Union’s Horizon 2020 research and innovation program (NF, NH: grant number 682540 – TransMID project; CF, NH: grant number 101003688 – EpiPose project).

## Conflict of interest

none

## Funding statement

The authors received no financial support for this research

## Notes

### Competing Interest Statement

The authors have declared no competing interest.

### Funding Statement

The authors received no financial support for this research. SD is supported by the Fonds National de la Recherche Scientifique (FNRS, Belgium). NF, CF and NH acknowledge support from the European Union's Horizon 2020 research and innovation program (NF, NH: grant number 682540 - TransMID project; CF, NH: grant number 101003688 - EpiPose project).

### Author Declarations

Ethics comittee of Saint-Pierre University Hospital (Brussels, Belgium)

## References

1. Vandenberg, O., Martiny, D., Rochas, O., van Belkum, A. & Kozlakidis, Z. Considerations for diagnostic COVID-19 tests. Nat. Rev. Microbiol. 19, 171–183 (2021).

2. Van den Wijngaert, S. et al. Bigger and Better? Representativeness of the Influenza A Surveillance Using One Consolidated Clinical Microbiology Laboratory Data Set as Compared to the Belgian Sentinel Network of Laboratories. Front. Public Health 7, 150 (2019).

3. Vandenberg, O. et al. Consolidation of Clinical Microbiology Laboratories and Introduction of Transformative Technologies. Clin. Microbiol. Rev. 33, (2020).

4. Albiger, B., Revez, J., Leitmeyer, K. C. & Struelens, M. J. Networking of Public Health Microbiology Laboratories Bolsters Europe’s Defenses against Infectious Diseases. Front. Public Health 6, (2018).

5. Vandenberg, O., Kozlakidis, Z., Schrenzel, J., Struelens, M. J. & Breuer, J. Control of Infectious Diseases in the Era of European Clinical Microbiology Laboratory Consolidation: New Challenges and Opportunities for the Patient and for Public Health Surveillance. Front. Med. 5, 15 (2018).

6. Van Oyen, H. & De Cock, J. Obligation de rapportage des résultats PCR/antigéniques et sérologiques dans le cadre de la pandémie COVID-19. (2020).

7. Cori, A., Ferguson, N. M., Fraser, C. & Cauchemez, S. A New Framework and Software to Estimate Time-Varying Reproduction Numbers During Epidemics. Am. J. Epidemiol. 178, 1505–1512 (2013).

8. Yin, N. et al. SARS-CoV-2 Diagnostic Tests: Algorithm and Field Evaluation From the Near Patient Testing to the Automated Diagnostic Platform. Front. Med. 8, 650581 (2021).

9. Hay, J. A. et al. Estimating epidemiologic dynamics from cross-sectional viral load distributions. MedRxiv Prepr. Serv. Health Sci. (2021) doi:10.1101/2020.10.08.20204222.

10. Risk Assessment Group. RAG Interpretation and Reporting of SARS-CoV-2 PCR Results. (2020).

11. Rasmussen, S. A. & Jamieson, D. J. Public Health Decision Making during Covid-19 — Fulfilling the CDC Pledge to the American People. N. Engl. J. Med. 383, 901–903 (2020).

12. Willem, L. et al. The impact of contact tracing and household bubbles on deconfinement strategies for COVID-19. Nat. Commun. 12, (2021).

13. Abrams, S. et al. Modelling the early phase of the Belgian COVID-19 epidemic using a stochastic compartmental model and studying its implied future trajectories. Epidemics 35, 100449 (2021).

14. Coletti, P. et al. A data-driven metapopulation model for the Belgian COVID-19 epidemic: assessing the impact of lockdown and exit strategies. BMC Infect. Dis. 21, (2021).

15. Franco, N. Covid-19 Belgium: Extended SEIR-QD model with nursing homes and long-term scenarios-based forecasts. medRxiv 2020.09.07.20190108 (2020) doi:10.1101/2020.09.07.20190108.

16. Hens, N., Faes, C. & Gilbert, M. On the timing of interventions to preserve hospital capacity: lessons to be learned from the Belgian SARS-CoV-2 pandemic. medRxiv 2020.12.18.20248450 (2020) doi:10.1101/2020.12.18.20248450.

17. Fowler, A. J. et al. Resource requirements for reintroducing elective surgery during the COVID-19 pandemic: modelling study. Br. J. Surg. 108, 97–103 (2021).

18. Frampton, D. et al. Genomic characteristics and clinical effect of the emergent SARS-CoV-2 B.1.1.7 lineage in London, UK: a whole-genome sequencing and hospital-based cohort study. Lancet Infect. Dis. (2021) doi:10.1016/S1473-3099(21)00170-5.

19. Muyldermans, G. et al. Surveillance of Infectious Diseases by the Sentinel Laboratory Network in Belgium: 30 Years of Continuous Improvement. PLOS ONE 11, e0160429 (2016).

20. Walckiers, D., Stroobant, A., Yourassowsky, E., Lion, J. & Cornelis, R. A sentinel network of microbiological laboratories as a tool for surveillance of infectious diseases in Belgium. Epidemiol. Infect. 106, 297–303 (1991).

21. Weemaes, M. et al. Laboratory information system requirements to manage the COVID-19 pandemic: A report from the Belgian national reference testing center. J. Am. Med. Inform. Assoc. 27, 1293–1299 (2020).

22. Harvala, H. et al. Emergence of a novel subclade of influenza A(H3N2) virus in London, December 2016 to January 2017. Eurosurveillance 22, (2017).

23. Singanayagam, A. et al. Duration of infectiousness and correlation with RT-PCR cycle threshold values in cases of COVID-19, England, January to May 2020. Eurosurveillance 25, (2020).

24. Dahdouh, E., Lázaro-Perona, F., Romero-Gómez, M. P., Mingorance, J. & García-Rodriguez, J. Ct values from SARS-CoV-2 diagnostic PCR assays should not be used as direct estimates of viral load. J. Infect. 82, 414–451 (2021).

25. Cornelissen, L. & André, E. Understanding the drivers of transmission of SARS-CoV-2. Lancet Infect. Dis. 21, 580–581 (2021).

26. Tom, M. R. & Mina, M. J. To Interpret the SARS-CoV-2 Test, Consider the Cycle Threshold Value. Clin. Infect. Dis. Off. Publ. Infect. Dis. Soc. Am. 71, 2252–2254 (2020).

